# Health concern affects egg consumption among older adults in Tamale Metropolis, Northern Ghana

**DOI:** 10.1101/2020.06.26.20140970

**Authors:** Humphrey Garti, Anthony Wemakor, Emmanuella Akwalpua, Nawaf Saeed

## Abstract

**Background:** Eggs are nutrient dense and rich source of quality protein and their consumption could significantly reduce incidence of chronic and infectious diseases especially in the elderly. This study determined egg consumption and factors that influence consumption among the elderly in Tamale metropolis.

**Methods:** Analytical cross-sectional study was conducted among 387 older adults 60 years and above. Data on egg consumption, nutritional knowledge and awareness of health implications of egg intake were collected using structured questionnaire. Relationships between egg consumption, nutritional knowledge and awareness of health effects of egg consumption were determined in bivariate and multivariate analysis.

**Results:** Respondents without awareness that egg intake increases risk of diseases were 10 times more likely to eat eggs compared to those who had awareness [Adjusted Odds Ratio 10.24; 95% Confidence Interval (CI) 4.20 – 25.00; p= 0.001]. Respondents with awareness that egg consumption was bad for health were less likely to consume eggs compared to those who thought egg intake was good for health [AOR 0.02; 95 % CI, 0.01 – 0.05, p < 0.001].

**Conclusion:** Egg consumption was low among the study population and was affected by health concerns: awareness that egg consumption was not good for health and exposes them to the risk of certain diseases

## INTRODUCTION

### Background

The proportion of persons aged 60 years and above in the Ghanaian population in 2000 was 7.2 % and is expected to reach 14.1 % by 2050 ^1^. The older adults are nutritionally vulnerable and exposed to increased risk of chronic diseases, disability and death. A number of factors (i.e., physiological changes, economic and social factors) significantly influence dietary intake in older adults ^2^. Inadequate nutrition, particularly reduced protein intake in older adults, is a significant contributor to sarcopenia, a predictor of early mortality and morbidities ^3,4^. Increased protein intake could be a useful strategy to promote good health and rapid recovery from illness in the elderly and reduce the incidence of sarcopenia among aging people ^3,5,6^.

Eggs provide rich natural source of high quality protein and a number of vital nutrients including vitamins A, B12, B5, and D, folate, choline, phosphorus, selenium, and good amounts of vitamins E, K, and B6, calcium and zinc ^7,8^. Egg has the highest value for protein digestibility (97 %) compared to all other sources of good quality protein ^9^ making it uniquely important for muscle protein synthesis than any other dietary protein ^5^. In relation to other sources of protein, eggs are inexpensive, easy to cook, chew and swallow and have long shelf life ^3^ and could be important source of protein for old people should it form integral part of their diets. Despite being nutritionally dense, eggs have been reported to increase the risk of certain lifestyles diseases by increasing blood cholesterol level ^10,11^ but this finding has not been supported by current research ^12^.

A study conducted in the United Kingdom suggested that older British adults consume 16 to 17 eggs a month with a reported protein intake of 1.24 ± 0.42g/kg body weight per day. In Ghana, there is hardly any scientific data on egg consumption for that matter protein intake by older adults. Such data could be necessary for development of effective national strategy to promote good quality protein intake among older adults and reduce age-related complications for improved health. This study therefore sought to determine the prevalence of egg consumption and its associated factors among older adults in Tamale Metropolis, Ghana.

### Methods

#### Study Area

This study was conducted in Tamale Metropolis of Northern Region of Ghana. The Metropolis has estimated land size of 646.90 km^2^ and a total population of 233,252. It is boarded by Sagnarigu district to the west and north, Mion district to the east, East Gonja district to the south and Central Gonja to the south west ^13^. The persons aged 60 years and above constitute 5.1 % of the population of the metropolis ^13^. The metropolis has one rainy season in a year from May to October, and a long dry season characterized by dry harmattan winds from November to February.

#### Study design, population and sampling

A cross-sectional study was conducted between April and May, 2018 among respondents aged 60 years and above in ten selected urban and peri-urban communities: Sawaba, Gunayeli, Aboabo, Dungu-kukuo, Choggu, Bilpela, Nyohini, Kunyevilla, Fuo-yepala and Wombele,

Sample size was determined using the formula N = [Z^2^p (1-p)]/ME^2 14^, where N = sample size, Z = confidence interval (CI) at 95 % =1.96, p = assumed prevalence of egg consumption among older adults (50 %), q = 1-0.5 = 0.5, ME= margin of error (5% = 0. 05). This yielded 384 so a sample size of 387 was used.

Systematic random sampling was used in selecting the participants of the study. Whenever a selected house had more than one eligible respondent, only one was selected using simple random sampling.

#### Inclusion and exclusion criteria

An older adult was eligible for interview if he or she was resident in a selected community, was able to answer questions and consented to be part of the study, or excluded if otherwise.

#### Data Collection

Data were collected in face-to-face interviews in homes using a pre-tested structured questionnaire. Data were collected on socio-demographic and economic characteristics of respondents (e.g., education, marital status, religion, income), and egg consumption (i.e., frequency of egg consumption, perception of disease risk associated with egg consumption). The questions on egg consumption were largely drawn from studies similar to ours ^15,16^ and used without further statistical validation.

#### Quality Control

The questionnaire was pre-tested in two communities outside the selected ones and any ambiguous questions restructured. For example, questions that elicited implausible responses (i.e., responses that were not meaningful and/or related to possible responses) were deemed to be ambiguous. Such questions were restructured with the help of language experts to ensure they conveyed the right meanings. The data collectors were taken through the questionnaire for common understanding and administration. The administered questionnaires were checked on the field for completeness and consistency by supervisors at the end of each day before the teams left the communities.

#### Data Analysis

Data management and analysis were done using Statistical Package for Social Sciences (SPSS) (version 21) for windows. Frequencies and percentages were computed for categorical data. Associations between consumption of eggs (dependent variable) and associated factors (independent variables) were explored in bivariate and multivariate analyses. Bivariate analysis was done as first-line test using chi-square to identify possible predictors of egg consumption. Factors with significant associations at P = 0.025 17,18were included in a logistic regression analysis to determine their independent effects. In the final model statistical significance was declared at p<0.05.

### Results

#### Socio-demographic characteristics of respondents

Majority (71.8 %) of the respondents were within 60-69 years age group, 86.8 % were Dagombas, and 98.7 % were Moslems. Majority (53.0 %) were males whilst 59.2 % and 52.2 % were self-employed and lived in rural areas of the metropolis respectively (Table 1). Majority, 76.2 % of the respondents, had no formal education and 79.8% were married.

**Table 1:** Socio-demographic characteristics of respondents (n=387)

| **Variable** | **Frequency (n)** | **Percent (%)** |
| --- | --- | --- |
| **Age group (years)** |  |  |
| 60-69 | 278 | 71.8 |
| 70 and above | 109 | 28.2 |
| **Sex** |  |  |
| Male | 205 | 53.0 |
| Female | 182 | 47.0 |
| **Religion** |  |  |
| Christianity | 40 | 10.3 |
| Islam | 347 | 89.7 |
| **Ethnicity** |  |  |
| Dagomba | 336 | 86.8 |
| Gonja | 26 | 6.7 |
| Mamprusi | 8 | 2.1 |
| Akan | 11 | 2.8 |
| Others | 6 | 1.6 |
| **Highest educational level** |  |  |
| None | 295 | 76.2 |
| Primary | 54 | 14.0 |
| Junior High /Middle School | 23 | 5.9 |
| Senior High School | 7 | 1.8 |
| Tertiary | 8 | 2.1 |
| **Marital status** |  |  |
| Single | 1 | 0.3 |
| Married | 309 | 79.8 |
| Divorced | 6 | 1.6 |
| Widowed | 69 | 17.8 |
| Separated | 2 | 0.5 |
| **Employment status** |  |  |
| Salaried worker | 10 | 2.6 |
| Self employed | 229 | 59.2 |
| Unemployed | 132 | 34.1 |
| Retired | 16 | 4.1 |
| **Residential area** |  |  |
| Urban | 39 | 10.1 |
| Rural | 202 | 52.2 |
| Peri-Urban | 146 | 37.7 |

#### Egg consumption knowledge of respondents

Results of the respondents’ knowledge and perception on egg as a constituent of the diet are presented in Table 2. Most (81.7 %) of the respondents consumed eggs but the rest claimed they did not consume it because of certain traditional beliefs and practices. Whilst most (71.8 %) thought egg consumption was good for health, 18.1 % thought it exposed them to increased risk of certain lifestyle diseases for which reason 64.3 % of these people did not eat it. About 26 % of the respondents mentioned at least one non-communicable disease attributed to the consumption of eggs.

**Table 2:** Egg consumption practices and knowledge of respondents (n=387)

| Variable | Frequency (n) | Percent (%) |
| --- | --- | --- |
| Do you consume eggs? |  |  |
| Yes | 316 | 81.7 |
| No | 71 | 18.3 |
| Do you think eggs are good for your health? |  |  |
| Yes | 278 | 71.8 |
| No | 57 | 14.7 |
| Don't know | 52 | 13.4 |
| Do you think the consumption of eggs increases your risk of certain lifestyle diseases? |  |  |
| Yes | 70 | 18.1 |
| No | 257 | 66.4 |
| Don't know | 60 | 15.5 |
| If yes, does that prevent you from egg consumption? |  |  |
| Yes | 45 | 64.3 |
| No | 25 | 35.7 |
| Are you forbidden from eating eggs? |  |  |
| Yes | 71 | 18.3 |
| No | 316 | 81.7 |
| Mentioned at least one disease associated with egg consumption |  |  |
| Yes | 102 | 26.4 |
| No | 285 | 73.6 |
| Have you been advised against the consumption of eggs by a health practitioner? |  |  |
| No | 387 | 100.0 |
| Yes | 0 | 0.0 |

#### Egg consumption and its associated factors

Majority (70.0%) of the respondents ate 10 eggs a month. Majority (64.2 %) of the respondents kept domestic birds with 65.5 % of those who kept birds consuming eggs from their birds. While most of the respondents (49.4 %) were unable to quantify their daily expenditure on eggs, 41.8 % spent not more than one Ghana Cedis (GHS1.00) (GHS5.00 = USD 1) a day on it (Table 3). The factors reported to influence egg intake by the respondents were availability (38.0 %) and nutrient content of eggs or health concern (26.6 %). Majority (54.1 %) ate eggs occasionally and at most one egg at a time (58.5 %). Two weeks prior to the study, more than 90 % of the respondents went at least a day without eating egg due to lack of money and this happened more than 6 times a week for the majority (53.6 %) of them.

**Table 3:** Egg consumption practices of egg consumers (n=316)

| **Variable** | **Frequency (n)** | **Percent (%)** |
| --- | --- | --- |
| What influences your egg intake? |  |  |
| Taste | 61 | 19.3 |
| Cost | 51 | 16.1 |
| Availability | 120 | 38.0 |
| Nutrient content/health concern | 84 | 26.6 |
| How often do you eat eggs in a week? |  |  |
| 1 time | 44 | 13.9 |
| 2 – 3 times | 56 | 17.7 |
| 4 – 5 times | 30 | 9.5 |
| 6 – 7 times | 15 | 4.7 |
| Occasionally | 171 | 54.1 |
| How many eggs do you eat at a time? |  |  |
| 1 | 185 | 58.5 |
| 2 | 128 | 40.5 |
| 3 | 3 | 0.9 |
| Do you keep birds? |  |  |
| Yes | 203 | 64.2 |
| No | 113 | 35.8 |
| Which of these birds do you keep? |  |  |
| Chickens | 111 | 35.1 |
| Guinea Fowls | 90 | 28.5 |
| Ducks | 1 | 0.3 |
| Others | 1 | 0.3 |
| Do you consume the eggs laid by the birds you keep? |  |  |
| Yes | 133 | 65.5 |
| No | 70 | 34.5 |
| How much do you spend on eggs a day? |  |  |
| 1 GHS *or less | 132 | 41.8 |
| 2 – 3 GHS* | 27 | 8.5 |
| 3 GHS* or more | 1 | 0.3 |
| I can't quantify | 156 | 49.4 |
| In the last two weeks has there been a day you ate food without eggs because there was no money to buy it? |  |  |
| Yes | 293 | 92.7 |
| No | 23 | 7.3 |
| How often did this happen? |  |  |
| Rarely (1-2 times) | 51 | 17.4 |
| Often (3-6 times) | 85 | 29.0 |
| Most Often (more than 6 times) | 157 | 53.6 |

\*GHS5 = USD 1

Possible factors associated with egg consumption measured in this study were tested in bivariate analysis for statistical association. Education, monthly income, eggs being forbidden, perception that eggs are bad for health, perception that egg consumption increases risk of certain diseases and area of residence were significantly associated with egg consumption in bivariate analyses (Table 4).

**Table 4:** Cross-tabulation of socio-demographic characteristics, knowledge on eggs and egg consumption of respondents.

| Characteristic | Egg consumption |  | Total | Test statistic |
| --- | --- | --- | --- | --- |
| Age group (years) | Yes (%) | No (%) |  |  |
| 60-69 | 231 (83.1) | 47 (16.9) | 278 | $\chi^2=1.37$;<br>$p=0.24$ |
| 70 and above | 85 (78.0) | 24 (22.0) | 109 |  |
| **Sex** |  |  |  |  |
| Male | 165 (80.5) | 40 (19.5) | 205 | $\chi^2=0.40$;<br>$p=0.50$ |
| Female | 151 (83.0) | 31 (17.0) | 182 |  |
| **Marital Status** |  |  |  |  |
| Married | 252 (81.6) | 71 (18.4) | 309 | $\chi^2=0.01$;<br>$p=0.92$ |
| Not currently married | 64 (82.1) | 14 (17.9) | 78 |  |
| **Religion** |  |  |  |  |
| Islam | 283 (81.6) | 64 (18.4) | 347 | $\chi^2=0.02$;<br>$p=0.88$ |
| Christian | 33 (82.5) | 7 (17.5) | 40 |  |
| **Ethnicity** |  |  |  |  |
| Dagomba | 275 (81.8) | 61 (18.2) | 336 | $\chi^2=0.49$;<br>$p=0.78$ |
| Gonja | 20 (76.9) | 6 (23.1) | 26 |  |
| Others | 21 (84.0) | 4 (16) | 25 |  |
| **Residential area** |  |  |  |  |
| Urban | 27 (69.2) | 12 (30.8) | 39 | $\chi^2=8.61$;<br>$p=0.01$ |
| Rural | 175 (86.6) | 27 (13.4) | 202 |  |
| Peri-urban | 114 (78.1) | 32 (21.9) | 146 |  |
| **Education** |  |  |  |  |
| Not educated | 235 (79.7) | 60 (20.3) | 295 | $\chi^2=3.29$;<br>$p=0.07$ |
| Educated | 81 (88.0) | 11 (12.0) | 92 |  |
| **Monthly income** |  |  |  |  |
| Less than GHS400* | 209 (84.6) | 38 (15.4) | 247 | $\chi^2=5.80$;<br>$p=0.07$ |
| GHS400* and above | 33 (82.5) | 7 (17.5) | 40 |  |
| Can't quantify | 74 (74.0) | 26 (26.0) | 100 |  |
| **Do you forbid eggs?** |  |  |  |  |
| Yes | 71 (100.0) | 0 (0.0) | 71 | $\chi^2=387.00$; |
| No | 0 (0.0) | 316 (100.0) | 316 | p<0.001 |
| **What effect (s) has egg consumption on your health?** |  |  |  |  |
| Good | 273 (98.2) | 5 (1.8) | 278 | $\chi^2=180.418$;<br>p<0.001 |
| Bad | 43 (39.4) | 66 (60.6) | 109 |  |
| **Do you think egg consumption increases your risk of certain diseases?** |  |  |  |  |
| Yes | 26 (37.1) | 44 (62.9) | 70 | $\chi^2=113.02$;<br>p<0.001 |
| No | 290 (91.5) | 27 (8.5) | 317 |  |
| **Do you keep birds?** |  |  |  |  |
| Yes | 203 (84.2) | 38 (15.8) | 241 | $\chi^2=2.84$;<br>p=0.092 |
| No | 113 (77.4) | 33 (22.6) | 146 |  |

\*GHS5 = USD 1

Independent effects of these predictors were tested in logistic regression analysis for significance. Respondents who perceived that egg consumption was bad for health were less likely to consume it compared to those who did not have this perception [Adjusted Odds Ratio (AOR) 0.02; 95 % Confidence Interval (CI) 0.01 – 0.05; p < 0.001] (Table 5). Similarly, those without the perception that egg consumption increases the risk of certain diseases were 10 times more likely to consume it compared to those with this perception (AOR 10.24; 95% CI 4.20 – 25.00; p= 0.001).

**Table 5:** Determinants of egg consumption.

| **Characteristic** | **Adjusted OR** | **95% Confidence Interval** | **P-value** |
| --- | --- | --- | --- |
| **Highest educational level** |  |  |  |
| No education | 0.56 | 0.16-1.98 | 0.370 |
| Some education | 1.00 |  |  |
| **Monthly income (GHS)\*** |  |  |  |
| Less than 400 | 2.38 | 0.49-11.50 | 0.280 |
| 400 and above | 1.00 |  |  |
| Can't quantify | 2.36 | 0.46-12.02 | 0.300 |
| **Residential area** |  |  |  |
| Urban | 1.00 |  |  |
| Rural | 1.88 | 0.52-6.74 | 0.335 |
| Peri-urban | 2.07 | 0.62-6.85 | 0.234 |
| **Do you keep birds?** |  |  |  |
| Yes | 1.00 |  |  |
| No | 1.31 | 0.57-3.02 | 0.532 |
| **Do you think the consumption of eggs increases your risk of certain diseases?** |  |  |  |
| Yes | 1.00 |  |  |
| No | 10.24 | 4.20-25.00 | <0.001 |
| **What effect(s) has egg consumption on your health?** |  |  |  |
| Good | 1.00 |  |  |
| Bad | 0.02 | 0.01-0.05 | <0.001 |

\*GHS5 = USD 1

### Discussion

This study sought to determine the prevalence of egg consumption and the factors that affect it in older adults in Tamale metropolis, Ghana. The findings of the study revealed that the prevalence of egg consumption in the older adults was low, and was mainly affected by the perception that egg consumption has adverse health effects and increases the risk of certain lifestyle diseases.

The study subjects consumed 10 eggs a month but we did not find any suitable studies on Ghanaian older adults for comparison. However, the prevalence of egg consumption measured compares favourably to an estimate of about 12 eggs a month reported for Ghanaians in 2017 ^19^. It was also consistent with consumption in South Africa where consumption per person per year was 150 eggs ^20^. Egg consumption was rather very low in many African countries. For instance in Zambia, Namibia and Botswana, consumption was 4, 8 and approximately 3 eggs per person per month respectively ^20^. Comparatively egg consumption in Africa was much lower than the reported consumption of 22 and 16-17 eggs a month among British adults 55 years and above, and 65 years and above respectively ^21^. Though the proportion of the study participants who admitted consuming egg was high (81.7 %), consumption was mostly occasional and at those instances only one egg was eaten at a time.

From this study the main determinant of egg consumption among older adults in Tamale metropolis was health concern. The perceptions that egg consumption was not good for health and it exposes one to risk of certain diseases showed strong association with egg consumption. Individuals who had the perception that egg consumption had adverse health effects and could specifically increase the risk of certain lifestyle diseases were less likely to consume it. This fear was probably founded on earlier publications which associated high cholesterol containing foods to high blood cholesterol level and coronary heart disease ^10^. A prospective cohort study involving a large cohort of physicians in the United States strongly associated consumption of 7 or more eggs a week with greater risks of cardiovascular disease and death ^11^. However, results from a meta-analysis of prospective cohort studies with large sample sizes and long durations of follow-up showed that an egg a day is safe and is not associated with an increased risk of cardiovascular disease or stroke ^12^. Blood cholesterol level attributable to consumption of eggs presents no significant health risk than cholesterol derived from consumption of dietary saturated fatty acids ^22,23^. Based on epidemiological evidence ^24^ egg could be safely consumed by everybody including those diagnosed of cardiovascular disease or even type II diabetes in whom mortality was earlier strongly associated with egg consumption ^11^. Though these findings eased restrictions on egg intake, there was still the fear among older adults in the current study who may be reluctant to change their eating habits to accommodate eggs in order to maintain low blood cholesterol ^3^. Even though area of residence, education, monthly income, and traditional practices and beliefs (forbid/taboo egg) strongly associated with egg consumption at bivariate level, they were not statistically significant at multivariate level.

Inadequate protein intake among older adults is a major contributor to sarcopenia and chronic diseases ^6^. Poor dentition, inability to chew and dysphagia reduce food intake in older adults ^3^ and contribute to inadequate protein intake ^25,26^. An increased intake of eggs could help reduce protein deficiency and associated health complications in older adults.

### Conclusion

This study shows that the prevalence of egg consumption is low among older adults and is affected by the perception it increases the risk of certain diseases. Health promotion efforts aimed at promoting egg consumption in older adults should address health concerns.

## LIMITATION

The study may suffer from recall bias since the egg consumption information was self-reported. Cross-sectional nature of the study prevented drawing of causal inference.

Our results are generalizable to older adult population in Tamale Metropolis.

## Data Availability

Dataset relating to this article are available and could be obtained from the corresponding author upon reasonable request.

## List of abbreviations

AOR: Adjusted odds ratio
CI: Confidence interval
LCPUFA: Long chain polyunsaturated fatty acid
GHS: Ghana cedis
SPSS: Statistical Package for Social Sciences

## DECLARATIONS

### Ethics approval and consent to participate

The study protocol was approved by the Joint Ethical Review Committee of School of Medicine and Health Sciences and School of Allied Health Sciences, University for Development Studies, Tamale, Ghana (Protocol Number 01-2018). The study objectives were carefully explained to the participants who were also informed of privacy and data protection and written informed consent was obtained from each of them before recruitment onto the project. They were also informed of opportunity to withdraw from the study at any time if they so wished.

### Consent for publication

Not applicable.

## Acknowledgement

Authors thank the aged for their time in answering questions and also for making themselves available for measurements to be taken on them. We are thankful to Patience Boanyah and Salome Vida Lieru for helping in data collection.

## Authors’ contribution

HG designed the study and supervised data collection. AW analysed and interpreted data. EA and NS collected data from field and drafted initial manuscript. HG and AW reviewed the draft manuscript. All authors read and approved the final manuscript.

## References

(1) Mba, C. J. Population ageing in Ghana: research gaps and the way forward. Journal of aging research 2010, 2010.

(2) Hewitt, G., Ismail, S.; Patterson, S.; Draper, A. The nutritional vulnerability of older Guyanese in residential homes. West Indian Medical Journal 2006, 55, 334–339.

(3) Smith, A.; Gray, J. Considering the benefits of egg consumption for older people at risk of sarcopenia. British journal of community nursing 2016, 21, 305–309.

(4) Morley, J. E.; Anker, S. D.; von Haehling, S.: Prevalence, incidence, and clinical impact of sarcopenia: facts, numbers, and epidemiology—update 2014. Springer, 2014.

(5) Bauer, J.; Biolo, G.; Cederholm, T.; Cesari, M.; Cruz-Jentoft, A. J.; Morley, J. E.; Phillips, S.; Sieber, C.; Stehle, P.; Teta, D. Evidence-based recommendations for optimal dietary protein intake in older people: a position paper from the PROT-AGE Study Group. Journal of the american Medical Directors association 2013, 14, 542–559.

(6) Murton, A. J. Muscle protein turnover in the elderly and its potential contribution to the development of sarcopenia. Proceedings of the Nutrition Society 2015, 74, 387–396.

(7) Ruxton, C.; Derbyshire, E.; Gibson, S. The nutritional properties and health benefits of eggs. Nutrition & Food Science 2010, 40, 263–279.

(8) Layman, D. K.; Rodriguez, N. R. Egg protein as a source of power, strength, and energy. Nutrition Today 2009, 44, 43–48.

(9) Organization, W. H., University, U. N.: Protein and amino acid requirements in human nutrition; World Health Organization, 2007; Vol. 935.

(10) Kannel, W.; Castelli, W.; McNamara, P. M. Serum lipid fractions and risk of coronary heart disease. The Framingham study. Minnesota medicine 1969, 52, 1225–1230.

(11) Djoussé, L.; Gaziano, J. M. Egg consumption in relation to cardiovascular disease and mortality: the Physicians’ Health Study–. The American journal of clinical nutrition 2008, 87, 964–969.

(12) Rong, Y.; Chen, L.; Zhu, T.; Song, Y.; Yu, M.; Shan, Z.; Sands, A.; Hu, F. B.; Liu, L. Egg consumption and risk of coronary heart disease and stroke: doseresponse meta-analysis of prospective cohort studies. Bmj 2013, 346, e8539.

(13) Service, G. S.: 2010 population and housing census report; Ghana Statistical Service, 2014.

(14) Snedecor, G. W.; Cochran, W. G. Statistical methods, 8thEdn. Ames: Iowa State Univ. Press Iowa 1989.

(15) van den Heuvel, E.; Murphy, J.; Appleton, K. Exploring the Consumption of Eggs in Older Adults: a Questionnaire Study. Proceedings of the Nutrition Society 2016, 75.

(16) Van den Heuvel, E.; Murphy, J.; Appleton, K. Exploring the barriers and facilitators to the consumption of eggs and other protein rich foods using focus groups. Proceedings of the Nutrition Society 2015, 74.

(17) Hosmer Jr, D. W., Lemeshow, S.; Sturdivant, R. X.: Applied logistic regression; John Wiley & Sons, 2013; Vol. 398.

(18) Sitotaw, B.; Mekuriaw, H.; Damtie, D. Prevalence of intestinal parasitic infections and associated risk factors among Jawi primary school children, Jawi town, north-west Ethiopia. BMC infectious diseases 2019, 19, 341. 2017.

(19) Larnyoh, M. T. Research shows every Ghanaian ate 143 eggs in 2017.

(20) Guyonnet, V. Opportunities to grow the egg business in Africa. 2017.

(21) van den Heuvel, E.; Murphy, J.; Appleton, K. Increasing dietary protein intake in community dwelling older adults: protocol for a randomised controlled trial and baseline data. Proceedings of the Nutrition Society 2017, 76.

(22) Gray, J.; Griffin, B. Eggs and dietary cholesterol–dispelling the myth. Nutrition Bulletin 2009, 34, 66–70.

(23) Kanter, M. M.; Kris-Etherton, P. M.; Fernandez, M. L.; Vickers, K. C.; Katz, D. L. Exploring the factors that affect blood cholesterol and heart disease risk: is dietary cholesterol as bad for you as history leads us to believe? Advances in nutrition 2012, 3, 711–717.

(24) Fuller, N.; Sainsbury, A.; Caterson, I.; Markovic, T. Egg consumption and human cardio-metabolic health in people with and without diabetes. Nutrients 2015, 7, 7399–7420.

(25) WHO. Protein and amino acid requirements in human nutrition. World health organization technical report series 2007, 1.

(26) Tieland, M.; Borgonjen-Van den Berg, K. J.; van Loon, L. J.; de Groot, L. C. Dietary protein intake in community-dwelling, frail, and institutionalized elderly people: scope for improvement. European journal of nutrition 2012, 51, 173–179.

